# Test performance evaluation of SARS-CoV-2 serological assays

**DOI:** 10.1101/2020.04.25.20074856

**Authors:** Jeffrey D. Whitman, Joseph Hiatt, Cody T. Mowery, Brian R. Shy, Ruby Yu, Tori N. Yamamoto, Ujjwal Rathore, Gregory M. Goldgof, Caroline Whitty, Jonathan M. Woo, Antonia E. Gallman, Tyler E. Miller, Andrew G. Levine, David N. Nguyen, Sagar P. Bapat, Joanna Balcerek, Sophia A. Bylsma, Ana M. Lyons, Stacy Li, Allison Wai-yi Wong, Eva Mae Gillis-Buck, Zachary B. Steinhart, Youjin Lee, Ryan Apathy, Mitchell J. Lipke, Jennifer Anne Smith, Tina Zheng, Ian C. Boothby, Erin Isaza, Jackie Chan, Dante D. Acenas, Jinwoo Lee, Trisha A. Macrae, Than S. Kyaw, David Wu, Dianna L. Ng, Wei Gu, Vanessa A. York, Haig Alexander Eskandarian, Perri C. Callaway, Lakshmi Warrier, Mary E. Moreno, Justine Levan, Leonel Torres, Lila A. Farrington, Rita Loudermilk, Kanishka Koshal, Kelsey C. Zorn, Wilfredo F. Garcia-Beltran, Diane Yang, Michael G. Astudillo, Bradley E. Bernstein, Jeffrey A. Gelfand, Edward T. Ryan, Richelle C. Charles, A. John Iafrate, Jochen K. Lennerz, Steve Miller, Charles Y. Chiu, Susan L. Stramer, Michael R. Wilson, Aashish Manglik, Chun Jimmie Ye, Nevan J. Krogan, Mark S. Anderson, Jason G. Cyster, Joel D. Ernst, Alan H. B. Wu, Kara L. Lynch, Caryn Bern, Patrick D. Hsu, Alexander Marson

## Abstract

**Background:** Serological tests are crucial tools for assessments of SARS-CoV-2 exposure, infection and potential immunity. Their appropriate use and interpretation require accurate assay performance data.

**Method:** We conducted an evaluation of 10 lateral flow assays (LFAs) and two ELISAs to detect anti-SARS-CoV-2 antibodies. The specimen set comprised 128 plasma or serum samples from 79 symptomatic SARS-CoV-2 RT-PCR-positive individuals; 108 pre-COVID-19 negative controls; and 52 recent samples from individuals who underwent respiratory viral testing but were not diagnosed with Coronavirus Disease 2019 (COVID-19). Samples were blinded and LFA results were interpreted by two independent readers, using a standardized intensity scoring system.

**Results:** Among specimens from SARS-CoV-2 RT-PCR-positive individuals, the percent seropositive increased with time interval, peaking at 81.8-100.0% in samples taken >20 days after symptom onset. Test specificity ranged from 84.3-100.0% in pre-COVID-19 specimens. Specificity was higher when weak LFA bands were considered negative, but this decreased sensitivity. IgM detection was more variable than IgG, and detection was highest when IgM and IgG results were combined. Agreement between ELISAs and LFAs ranged from 75.7-94.8%. No consistent cross-reactivity was observed.

**Conclusion:** Our evaluation showed heterogeneous assay performance. Reader training is key to reliable LFA performance, and can be tailored for survey goals. Informed use of serology will require evaluations covering the full spectrum of SARS-CoV-2 infections, from asymptomatic and mild infection to severe disease, and later convalescence. Well-designed studies to elucidate the mechanisms and serological correlates of protective immunity will be crucial to guide rational clinical and public health policies.

## INTRODUCTION

As of May 11, 2020, more than 285,000 deaths have been attributed to Coronavirus Disease 2019 (COVID-19).^1^ Millions of infections by SARS-CoV-2, the virus responsible for COVID-19, have been reported, though its full extent has yet to be determined due to limited testing.^2^ Government interventions to slow viral spread have disrupted daily life and economic activity for billions of people. Strategies to ease restraints on human mobility and interaction, without provoking major resurgence of transmission and mortality, will depend on accurate estimates of population levels of infection and immunity.^3^ Current testing for the virus largely depends on labor-intensive molecular techniques.^4^ Individuals with positive molecular tests represent only a small fraction of all infections, given limited deployment and the brief time window when PCR testing has the highest sensitivity.^5-7^ The proportion of undocumented cases in the original epidemic focus was estimated to be as high as 86%,^8^ and asymptomatic infections are suspected to play a substantial role in transmission.^9-14^

Widely available, reliable antibody detection assays would enable more accurate estimates of SARS-CoV-2 prevalence and incidence. On February 4, 2020, the Secretary of the United States Department of Health and Human Services issued emergency use authorization (EUA) for diagnosis of SARS-CoV-2,^15^ allowing nucleic acid detection and immunoassay tests to be offered based on manufacturer-reported data without formal FDA clearance.^16^ In response, dozens of companies began to market laboratory-based immunoassays and point-of-care tests. Rigorous, comparative performance data are crucial to inform clinical care and public health responses.

We conducted a head-to-head comparison of serology tests available to our group – comprised of immunochromatographic lateral flow assays (LFAs) and enzyme-linked immunosorbent assays (ELISAs). Our evaluation includes performance by time from symptom onset and disease severity. Our goal is to provide well-controlled performance data to help guide their potential development and deployment.

## METHODS

### Ethical approvals

This study was approved by institutional review boards at the University of California, San Francisco (UCSF)/Zuckerberg San Francisco General Hospital (ZSFG) and Massachusetts General Hospital (MGH).

### Study Design

The study population included individuals with symptomatic infection and positive SARS-CoV-2 real-time polymerase chain reaction (RT-PCR) testing of nasopharyngeal or oropharyngeal swabs, who had remnant serum and plasma specimens in clinical laboratories serving the UCSF and ZSFG Medical Center networks. We included multiple specimens per individual, but no more than one sample per time interval (1-5, 6-10, 11-15, 16-20, and >20 days after symptom onset). If an individual had more than one specimen for a given time interval, only the later specimen was included. For specificity, we included 108 pre-COVID-19 plasma specimens from eligible blood donors collected prior to July 2018.^17^ We assessed cross-reactivity using 52 specimens from 2020: 50 with test results for other respiratory viruses (Biofire FilmArray; BioFire Diagnostics, Salt Lake City, UT), and 32 with negative results by SARS-CoV-2 RT-PCR. We based minimum sample size calculations on expected binomial exact 95% confidence limits. A total of 288 samples were included in the final analysis, including 128 from 79 SARS-CoV-2 RT-PCR-positive individuals. Some specimens were exhausted during the analysis and were not included in all tests. Data obtained from serial specimens that did not conform to our study design were excluded.

Clinical data were extracted from electronic health records and entered in a HIPAA-secure REDCap database hosted by UCSF. Data included demographic information, major co-morbidities, patient-reported symptom onset date, symptoms and indicators of severity.

Independent data from testing efforts at MGH, with slight deviations in methods, are included as Supplementary Data. Briefly, 57 heat-inactivated serum/plasma samples from 44 SARS-CoV-2 RT-PCR-positive individuals were included. For specificity, the MGH study included 60 heat-inactivated, pre-COVID-19 samples from 30 asymptomatic adults and 30 individuals admitted with febrile and/or respiratory illness with a confirmed pathogen.

### Sample Preparation

Samples from UCSF and ZSFG were assigned a random well position in one of four 96-well plates. Samples were thawed at 37°C, and up to 200uL was transferred to the assigned well without heat inactivation. Samples were then sub-aliquoted (12.5μL) to replica plates for testing. Replica plates were stored at −20°C until needed, then thawed for ten minutes at room temperature and briefly centrifuged before testing. All sample handling followed UCSF biosafety committee-approved practices.

For the MGH study, samples were heat-inactivated at 56°C for 60 minutes, aliquoted, and stored at 4°C and −20°C. Samples stored at 4°C were used within 7 days. Frozen aliquots were stored until needed with only a single freeze-thaw cycle for any sample. All samples were brought to room temperature and briefly centrifuged prior to adding the recommended volume to the LFA cartridge.

### Immunochromatographic Lateral Flow Assays (LFAs)

Ten lateral flow assays were evaluated (eTable 1). At the time of testing, cartridges were labeled by randomized sample location (plate, well). The appropriate sample volume was transferred from the plate to the indicated sample port, followed by provided diluent, following manufacturer instructions. The lateral flow cartridges were incubated for the recommended time at room temperature before readings. Each cartridge was assigned a semi-quantitative score (0 for negative, 1 to 6 for positive) for test line intensity by two independent readers blinded to specimen status and to each other’s scores (eFigure 1).^17^ For some cartridges (DeepBlue, UCP, Bioperfectus), the positive control indicator failed to appear after addition of diluent in a significant fraction of tests. For these tests, two further drops of diluent were added to successfully recover control indicators in all affected tests. These results were included in analyses. During testing, two plates were transposed 180° and assays were run in the opposite order from the wells documented on cartridges. These data were corrected and accuracy was confirmed by empty well position and verification of a subset of results.

### Enzyme-Linked Immunosorbent Assays (ELISAs)

Epitope Diagnostics ELISAs were performed according to manufacturer specifications. Cutoffs for IgG and IgM detection were calculated as the package insert described (see Supplementary Methods). Values greater than the cutoff were considered positive.

An in-house ELISA was performed with minor deviations from a published protocol.^18^ SARS-CoV-2 Receptor Binding Domain (RBD) protein was produced from the published construct (NR-52306, BEI Resources). The positive cutoff was equal to the mean of the OD values of the negative control wells on the respective plate plus three times the standard deviation of the OD value distribution from the 108 pre-COVID-19 plasma. For both ELISAs, background-corrected OD values were divided by the cutoff to generate signal-to-cutoff (S/CO) ratios. Samples with S/CO values greater than 1.0 were considered positive.

### Data Analysis

For LFA testing, the second reader’s scores were used for performance calculations, and the first reader’s score was used to calculate inter-reader agreement statistics. Percent seropositivity among RT-PCR-confirmed cases was calculated by time interval from symptom onset. Specificity was based on results in pre-COVID-2019 samples. Binomial exact 95% confidence intervals were calculated for all estimates. Analyses were conducted in R (3.6.3) and SAS (9.4).

## RESULTS

### Study population

SARS-CoV-2-positive individuals in the UCSF/ZSFG study ranged from 22 to >90 years of age (Table 1). The majority of SARS-CoV-2-positive individuals were Hispanic/Latinx (68%), reflecting the ZSFG patient population and demographics of the epidemic in San Francisco.^19,20^ Most presented with cough (91%) and fever (86%). Chronic medical conditions, such as hypertension, type 2 diabetes mellitus, obesity, and chronic kidney disease, were frequent. Of the 79 cases, 18% were outpatients, 46% inpatients without ICU care, and 37% required ICU care; there had been no reported deaths at the time of chart review.

**Table 1:** Baseline demographic characteristics, presenting symptoms, chronic medical conditions, initial disposition and highest-level outcome for all participants whose samples were included in each time interval for serological testing. Only one sample per patient was included in each time interval, and some individuals are represented by multiple samples in different time intervals. In total, we tested 128 samples taken from 79 SARS-CoV-2 RT-PCR-positive cases.

| Variable | All Patients<br>(N=79) | 0-5d<br>(N=28) | 6-10d<br>(N=36) | 11-15d<br>(N=34) | 16-20d<br>(N=19) | >20d<br>(N=11) |
| --- | --- | --- | --- | --- | --- | --- |
| Age, mean (S.D.) y | 52.9 (15) | 48.2 (15.0) | 53.3 (15.1) | 58.1±15.1 | 56.6 (13.2) | 55.5 (14.8) |
| Male sex (%) | 54 (68) | 15 (54) | 24 (67) | 21 (62) | 12 (63) | 8 (73) |
| **Racial or ethnic group** |  |  |  |  |  |  |
| Hispanic (%) | 54 (68) | 18 (64) | 29 (81) | 23 (68) | 12 (63) | 7 (64) |
| Asian (%) | 7 (9) | 3 (11) | 2 (6) | 4 (12) | 3 (16) | 0 (0) |
| White (%) | 7 (9) | 3 (11) | 1 (3) | 2 (6) | 2 (11) | 0 (0) |
| Black (%) | 6 (8) | 2 (7) | 3 (8) | 4 (12) | 1 (5) | 2 (18) |
| Other/not reported (%) | 5 (6) | 2 (7) | 1 (3) | 1 (3) | 1 (5) | 2 (18) |
| **Presenting symptoms** |  |  |  |  |  |  |
| Cough (%) | 72 (91) | 24 (86) | 33 (92) | 31 (91) | 17 (89) | 9 (82) |
| Fever (%) | 68 (86) | 23 (82) | 30 (83) | 29 (85) | 17 (89) | 9 (82) |
| Myalgia (%) | 29 (37) | 8 (29) | 12 (33) | 13 (38) | 8 (42) | 3 (27) |
| Chest pain (%) | 20 (25) | 5 (18) | 8 (22) | 7 (21) | 5 (26) | 4 (36) |
| Headache (%) | 20 (25) | 4 (14) | 11 (31) | 9 (26) | 6 (32) | 4 (36) |
| Chills (%) | 19 (24) | 5 (18) | 9 (25) | 7 (21) | 7 (37) | 2 (18) |
| Sore throat (%) | 19 (24) | 4 (14) | 11 (31) | 8 (24) | 5 (26) | 3 (27) |
| Malaise (%) | 17 (22) | 4 (14) | 7 (19) | 9 (26) | 4 (21) | 1 (9) |
| Diarrhea (%) | 13 (16) | 4 (14) | 7 (19) | 6 (18) | 4 (21) | 1 (9) |
| Anorexia (%) | 8 (10) | 2 (7) | 1 (3) | 2 (6) | 4 (21) | 1 (9) |
| Nausea and/or vomiting (%) | 8 (10) | 2 (7) | 2 (6) | 2 (6) | 2 (11) | 1 (9) |
| Anosmia and/or dysgeusia (%) | 4 (5) | 1 (4) | 1 (3) | 2 (6) | 0 (0) | 1 (9) |
| **Chronic medical conditions** |  |  |  |  |  |  |
| Hypertension (%) | 36 (46) | 11 (39) | 17 (47) | 21 (62) | 11 (58) | 6 (55) |
| T2DM (%) | 33 (42) | 11 (39) | 17 (47) | 19 (56) | 8 (42) | 6 (55) |
| Obesity (%) | 19 (24) | 7 (25) | 9 (25) | 11 (32) | 6 (32) | 6 (55) |
| CKD (%) | 10 (13) | 4 (14) | 3 (8) | 6 (18) | 4 (21) | 3 (27) |
| Hypothyroid (%) | 6 (8) | 3 (11) | 3 (8) | 3 (9) | 0 (0) | 0 (0) |
| Solid organ transplant (%) | 6 (8) | 2 (7) | 0 (0) | 2 (6) | 2 (11) | 2 (18) |
| CAD (%) | 5 (6) | 1 (4) | 1 (3) | 2 (6) | 2 (11) | 3 (27) |
| Asthma (%) | 4 (5) | 1 (4) | 1 (3) | 3 (9) | 2 (11) | 0 (0) |
| CHF (%) | 3 (4) | 2 (7) | 2 (6) | 2 (6) | 1 (5) | 0 (0) |
| Liver disease (%) | 3 (4) | 0 (0) | 1 (3) | 2 (6) | 1 (5) | 1 (9) |
| Malignancy (%) | 3 (4) | 1 (4) | 2 (6) | 1 (3) | 2 (11) | 0 (0) |
| Emphysema (%) | 2 (3) | 0 (0) | 1 (3) | 1 (3) | 1 (5) | 1 (9) |
| Prior stroke (%) | 2 (3) | 1 (4) | 0 (0) | 0 (0) | 0 (0) | 1 (9) |
| HIV (%) | 1 (1) | 1 (4) | 0 (0) | 0 (0) | 0 (0) | 0 (0) |
| Other immune compromised condition* (%) | 5 (6) | 1 (4) | 1 (3) | 3 (9) | 2 (11) | 1 (9) |
| **Highest-level of care** |  |  |  |  |  |  |
| Ambulatory** (%) | 14 (18) | 9 (32) | 2 (6) | 3 (9) | 0 (0) | 0 (0) |
| Admitted (%) | 36 (46) | 11 (39) | 19 (53) | 12 (35) | 5 (26) | 4 (36) |
| ICU (%) | 29 (37) | 8 (29) | 15 (42) | 19 (56) | 14 (74) | 7 (64) |

\*Other immune compromised condition includes rheumatology patients (rheumatoid arthritis, psoriasis, Crohn's disease, ankylosing spondylitis, and reactive arthritis), all of whom were taking immune modulating/suppressing therapies.
\*\*Ambulatory care includes outpatient as well as patients seen in ED and not admitted.

### Test Performance

The percentage of specimens testing positive rose with increasing time from symptom onset (Table 2, Figure 1A), reaching the highest levels in the 16-20 and >20 day time intervals. The highest detection rate was achieved by combining IgM and IgG results (Figure 1B). However, 95% confidence intervals for later time intervals showed substantial overlap with those for earlier intervals (Figure 1B). Four assays (Bioperfectus, Premier, Wondfo, in-house ELISA) achieved >80% positivity in the later two time intervals (16-20 and >20 days) while maintaining >95% specificity. Some tests were not performed on a subset of specimens due exhausted sample material, which may have affected reported percent positivity. IgM detection was less consistent than IgG for nearly all assays. Kappa agreement statistic ranged from 0.95 to 0.99 for IgG and 0.81-1.00 for IgM for standardized intensity score and training (eTable 2 and eFigure 2). Although variability in mean band intensities exists among different assays, the rate of sample positivity was generally consistent (Figure 2).

**Table 2:** Summary statistics for immunochromatographic lateral flow assays (LFAs) and Enzyme-Linked Immunosorbent Assays (ELISAs). Samples are binned by time after patient-reported symptom onset for SARS-CoV-2 RT-PCR-positive cases. Percent of seropositivity assessed by each assay in SARS-CoV-2 RT-PCR-positive samples is reported with 95% Confidence Intervals (95% CI). The column “IgM or IgG” refers to positivity of either isotype. Specificity is determined relative to pre-COVID-19 negative control serum samples. Percent of seropositivity assessed by each assay is reported with 95% confidence intervals for samples from individuals who were positive for non-SARS-CoV-2 viral infections and/or tested negative for SARS-CoV-2 by RT-PCR.

| Percentage of positive specimens from patients with positive SARS-CoV2 RT-PCR by days since symptom onset |  |  |  |  |  |  |  |  |  |  |  |  |
| --- | --- | --- | --- | --- | --- | --- | --- | --- | --- | --- | --- | --- |
| Assay | IgM |  |  |  | IgG |  |  |  | IgM or IgG |  |  |  |
|  | Total N | positive | % | 95% CI | Total N | positive | % | 95% CI | Total N | positive | % | 95% CI |
| Immunochromatographic Lateral Flow Assays |  |  |  |  |  |  |  |  |  |  |  |  |
| **Biomedomics** |  |  |  |  |  |  |  |  |  |  |  |  |
| 1-5 days | 27 | 7 | **25.9** | 11.1 - 46.3 | 27 | 6 | **22.2** | 8.6 - 42.3 | 27 | 8 | **29.6** | 13.8 - 50.2 |
| 6-10 days | 36 | 22 | **61.1** | 43.5 - 76.9 | 36 | 19 | **52.8** | 35.5 - 69.6 | 36 | 23 | **63.9** | 46.2 - 79.2 |
| 11-15 days | 33 | 25 | **75.8** | 57.7 - 88.9 | 33 | 23 | **69.7** | 51.3 - 84.4 | 33 | 26 | **78.8** | 61.1 - 91.0 |
| 16-20 days | 19 | 16 | **84.2** | 60.4 - 96.6 | 19 | 14 | **73.7** | 48.8 - 90.9 | 19 | 17 | **89.5** | 66.9 - 98.7 |
| >20 days | 11 | 9 | **81.8** | 48.2 - 97.7 | 11 | 9 | **81.8** | 48.2 - 97.7 | 11 | 9 | **81.8** | 48.2 - 97.7 |
| **Bioperfectus** |  |  |  |  |  |  |  |  |  |  |  |  |
| 1-5 days | 28 | 11 | **39.3** | 21.5 - 59.4 | 28 | 7 | **25.0** | 10.7 - 44.9 | 28 | 11 | **39.3** | 21.5 - 59.4 |
| 6-10 days | 35 | 26 | **74.3** | 56.7 - 87.5 | 35 | 23 | **65.7** | 47.8 - 80.9 | 35 | 27 | **77.1** | 59.9 - 89.6 |
| 11-15 days | 34 | 28 | **82.4** | 65.5 - 93.2 | 34 | 27 | **79.4** | 62.1 - 91.3 | 34 | 30 | **88.2** | 72.5 - 96.7 |
| 16-20 days | 19 | 16 | **84.2** | 60.4 - 96.6 | 19 | 14 | **73.7** | 48.8 - 90.9 | 19 | 17 | **89.5** | 66.9 - 98.7 |
| >20 days | 10 | 10 | **100.0** | 69.2 - 100.0 | 10 | 9 | **90.0** | 55.5 - 99.7 | 10 | 10 | **100.0** | 69.2 - 100.0 |
| **DecomBio** |  |  |  |  |  |  |  |  |  |  |  |  |
| 1-5 days | 26 | 8 | **30.8** | 14.3 - 51.8 | 26 | 7 | **26.9** | 11.6 - 47.8 | 26 | 8 | **30.8** | 14.3 - 51.8 |
| 6-10 days | 36 | 24 | **66.7** | 49.0 - 81.4 | 36 | 24 | **66.7** | 49.0 - 81.4 | 36 | 24 | **66.7** | 49.0 - 81.4 |
| 11-15 days | 33 | 29 | **87.9** | 71.8 - 96.6 | 33 | 29 | **87.9** | 71.8 - 96.6 | 33 | 29 | **87.9** | 71.8 - 96.6 |
| 16-20 days | 18 | 14 | **77.8** | 52.4 - 93.6 | 18 | 14 | **77.8** | 52.4 - 93.6 | 18 | 14 | **77.8** | 52.4 - 93.6 |
| >20 days | 11 | 10 | **90.9** | 58.7 - 99.8 | 11 | 10 | **90.9** | 58.7 - 99.8 | 11 | 10 | **90.9** | 58.7 - 99.8 |
| **DeepBlue** |  |  |  |  |  |  |  |  |  |  |  |  |
| 1-5 days | 28 | 12 | **42.9** | 24.5 - 62.8 | 28 | 6 | **21.4** | 8.3 - 41.0 | 28 | 12 | **42.9** | 24.5 - 62.8 |
| 6-10 days | 36 | 28 | **77.8** | 60.8 - 89.9 | 36 | 18 | **50.0** | 32.9 - 67.1 | 36 | 28 | **77.8** | 60.8 - 89.9 |
| 11-15 days | 34 | 28 | **82.4** | 65.5 - 93.2 | 34 | 21 | **61.8** | 43.6 - 77.8 | 34 | 28 | **82.4** | 65.5 - 93.2 |
| 16-20 days | 19 | 16 | **84.2** | 60.4 - 96.6 | 19 | 15 | **78.9** | 54.4 - 93.9 | 19 | 17 | **89.5** | 66.9 - 98.7 |
| >20 days | 11 | 10 | **90.9** | 58.7 - 99.8 | 11 | 9 | **81.8** | 48.2 - 97.7 | 11 | 10 | **90.9** | 58.7 - 99.8 |
| **Innovita** |  |  |  |  |  |  |  |  |  |  |  |  |
| 1-5 days | 27 | 4 | **14.8** | 4.2 - 33.7 | 27 | 7 | **25.9** | 11.1 - 46.3 | 27 | 7 | **25.9** | 11.1 - 46.3 |
| 6-10 days | 36 | 12 | **33.3** | 18.6 - 51.0 | 36 | 17 | **47.2** | 30.4 - 64.5 | 36 | 20 | **55.6** | 38.1 - 72.1 |
| 11-15 days | 31 | 12 | **38.7** | 21.8 - 57.8 | 32 | 25 | **78.1** | 60.0 - 90.7 | 32 | 25 | **78.1** | 60.0 - 90.7 |
| 16-20 days | 13 | 4 | **30.8** | 9.1 - 61.4 | 13 | 9 | **69.2** | 38.6 - 90.9 | 13 | 9 | **69.2** | 38.6 - 90.9 |
| >20 days | 6 | 1 | **16.7** | 0.4 - 64.1 | 6 | 4 | **66.7** | 22.3 - 95.7 | 6 | 5 | **83.3** | 35.9 - 99.6 |
| **Premier** |  |  |  |  |  |  |  |  |  |  |  |  |
| 1-5 days | 28 | 10 | **35.7** | 18.6 - 55.9 | 28 | 6 | **21.4** | 8.3 - 41.0 | 28 | 10 | **35.7** | 18.6 - 55.9 |
| 6-10 days | 35 | 25 | **71.4** | 53.7 - 85.4 | 35 | 18 | **51.4** | 34.0 - 68.6 | 35 | 25 | **71.4** | 53.7 - 85.4 |
| 11-15 days | 34 | 28 | **82.4** | 65.5 - 93.2 | 34 | 22 | **64.7** | 46.5 - 80.3 | 34 | 29 | **85.3** | 68.9 - 95.0 |
| 16-20 days | 19 | 16 | **84.2** | 60.4 - 96.6 | 19 | 14 | **73.7** | 48.8 - 90.9 | 19 | 17 | **89.5** | 66.9 - 98.7 |
| >20 days | 11 | 10 | **90.9** | 58.7 - 99.8 | 11 | 9 | **81.8** | 48.2 - 97.7 | 11 | 10 | **90.9** | 58.7 - 99.8 |
| **Sure** |  |  |  |  |  |  |  |  |  |  |  |  |
| 1-5 days | 28 | 3 | **10.7** | 2.3 - 28.2 | 28 | 5 | **17.9** | 6.1 - 36.9 | 28 | 5 | **17.9** | 6.1 - 36.9 |
| 6-10 days | 35 | 15 | **42.9** | 26.3 - 60.6 | 35 | 19 | **54.3** | 36.6 - 71.2 | 35 | 19 | **54.3** | 36.6 - 71.2 |
| 11-15 days | 34 | 22 | **64.7** | 46.5 - 80.3 | 34 | 25 | **73.5** | 55.6 - 87.1 | 34 | 25 | **73.5** | 55.6 - 87.1 |
| 16-20 days | 19 | 14 | **73.7** | 48.8 - 90.9 | 19 | 14 | **73.7** | 48.8 - 90.9 | 19 | 15 | **78.9** | 54.4 - 93.9 |
| >20 days | 11 | 8 | **72.7** | 39.0 - 94.0 | 11 | 10 | **90.9** | 58.7 - 99.8 | 11 | 10 | **90.9** | 58.7 - 99.8 |
| **UCP** |  |  |  |  |  |  |  |  |  |  |  |  |
| 1-5 days | 28 | 7 | **25.0** | 10.7 - 44.9 | 28 | 7 | **25.0** | 10.7 - 44.9 | 28 | 7 | **25.0** | 10.7 - 44.9 |
| 6-10 days | 36 | 21 | **58.3** | 40.8 - 74.5 | 36 | 18 | **50.0** | 32.9 - 67.1 | 36 | 21 | **58.3** | 40.8 - 74.5 |
| 11-15 days | 34 | 26 | **76.5** | 58.8 - 89.3 | 34 | 25 | **73.5** | 55.6 - 87.1 | 34 | 27 | **79.4** | 62.1 - 91.3 |
| 16-20 days | 19 | 15 | **78.9** | 54.4 - 93.9 | 19 | 14 | **73.7** | 48.8 - 90.9 | 19 | 15 | **78.9** | 54.4 - 93.9 |
| >20 days | 11 | 10 | **90.9** | 58.7 - 99.8 | 11 | 9 | **81.8** | 48.2 - 97.7 | 11 | 10 | **90.9** | 58.7 - 99.8 |
| **VivaChek** |  |  |  |  |  |  |  |  |  |  |  |  |
| 1-5 days | 25 | 7 | **28.0** | 12.1 - 49.4 | 25 | 7 | **28.0** | 12.1 - 49.4 | 25 | 7 | **28.0** | 12.1 - 49.4 |
| 6-10 days | 35 | 22 | **62.9** | 44.9 - 78.5 | 35 | 22 | **62.9** | 44.9 - 78.5 | 35 | 22 | **62.9** | 44.9 - 78.5 |
| 11-15 days | 30 | 26 | **86.7** | 69.3 - 96.2 | 30 | 25 | **83.3** | 65.3 - 94.4 | 30 | 26 | **86.7** | 69.3 - 96.2 |
| 16-20 days | 19 | 15 | **78.9** | 54.4 - 93.9 | 19 | 14 | **73.7** | 48.8 - 90.9 | 19 | 15 | **78.9** | 54.4 - 93.9 |
| >20 days | 10 | 9 | **90.0** | 55.5 - 99.7 | 10 | 9 | **90.0** | 55.5 - 99.7 | 10 | 9 | **90.0** | 55.5 - 99.7 |
| **WondFo** |  |  |  |  |  |  |  |  |  |  |  |  |
| 1-5 days |  |  |  |  |  |  |  |  | 26 | 10 | **38.5** | 20.2 - 59.4 |
| 6-10 days |  |  |  |  |  |  |  |  | 36 | 24 | **66.7** | 49.0 - 81.4 |
| 11-15 days |  |  |  |  |  |  |  |  | 32 | 27 | **84.4** | 67.2 - 94.7 |
| 16-20 days |  |  |  |  |  |  |  |  | 19 | 17 | **89.5** | 66.9 - 98.7 |
| >20 days |  |  |  |  |  |  |  |  | 11 | 9 | **81.8** | 48.2 - 97.7 |
| **ELISAs** |  |  |  |  |  |  |  |  |  |  |  |  |
| **Epitope** |  |  |  |  |  |  |  |  |  |  |  |  |
| 1-5 days | 28 | 5 | **17.9** | 6.1 - 36.9 | 28 | 11 | **39.3** | 21.5 - 59.4 | 28 | 11 | **39.3** | 39.3 - 21.5 |
| 6-10 days | 36 | 19 | **52.8** | 35.5 - 69.6 | 36 | 28 | **77.8** | 60.8 - 89.9 | 36 | 29 | **80.6** | 80.6 - 64.0 |
| 11-15 days | 34 | 27 | **79.4** | 62.1 - 91.3 | 34 | 31 | **91.2** | 76.3 - 98.1 | 34 | 31 | **91.2** | 91.2 - 76.3 |
| 16-20 days | 19 | 14 | **73.7** | 48.8 - 90.9 | 19 | 16 | **84.2** | 60.4 - 96.6 | 19 | 17 | **89.5** | 89.5 - 66.9 |
| >20 days | 11 | 9 | **81.8** | 48.2 - 97.7 | 11 | 10 | **90.9** | 58.7 - 99.8 | 11 | 10 | **90.9** | 90.9 - 58.7 |
| **In-House** |  |  |  |  |  |  |  |  |  |  |  |  |
| 1-5 days |  |  |  |  |  |  |  |  | 28 | 10 | **35.7** | 18.6 - 55.9 |
| 6-10 days |  |  |  |  |  |  |  |  | 36 | 26 | **72.2** | 54.8 - 85.8 |
| 11-15 days |  |  |  |  |  |  |  |  | 34 | 32 | **94.1** | 80.3 - 99.3 |
| 16-20 days |  |  |  |  |  |  |  |  | 19 | 17 | **89.5** | 66.9 - 98.7 |
| >20 days |  |  |  |  |  |  |  |  | 11 | 9 | **81.8** | 48.2 - 97.7 |
| **Specificity in 108 blood donor plasma specimens collected before July 2018** |  |  |  |  |  |  |  |  |  |  |  |  |
| **Assay** | **IgM** |  |  |  | **IgG** |  |  |  | **IgM or IgG** |  |  |  |
|  | **Total N** | **positive** | **%** | **95% CI** | **Total N** | **positive** | **%** | **95% CI** | **Total N** | **positive** | **%** | **95% CI** |
| **Immunochromatographic Lateral Flow Assays** |  |  |  |  |  |  |  |  |  |  |  |  |
| **Biomedomics** | 107 | 13 | **87.9** | 80.1 - 93.4 | 107 | 4 | **96.3** | 90.7 - 99.0 | 107 | 14 | **86.9** | 79.0 - 92.7 |
| **Bioperfectus** | 104 | 3 | **97.1** | 91.8 - 99.4 | 104 | 2 | **98.1** | 93.2 - 99.8 | 104 | 5 | **95.2** | 89.1 - 98.4 |
| **DecomBio** | 107 | 10 | **90.7** | 83.5 - 95.4 | 107 | 9 | **91.6** | 84.6 - 96.1 | 107 | 11 | **89.7** | 82.3 - 94.8 |
| **DeepBlue** | 108 | 17 | **84.3** | 76.0 - 90.6 | 108 | 1 | **99.1** | 94.9 - 100.0 | 108 | 17 | **84.3** | 76.0 - 90.6 |
| **Innovita** | 108 | 4 | **96.3** | 90.8 - 99.0 | 108 | 0 | **100.0** | 96.6 - 100.0 | 108 | 4 | **96.3** | 90.8 - 99.0 |
| **Premier** | 108 | 2 | **98.1** | 93.5 - 99.8 | 108 | 1 | **99.1** | 94.9 - 100.0 | 108 | 3 | **97.2** | 92.1 - 99.4 |
| **Sure** | 108 | 0 | **100.0** | 96.6 - 100.0 | 108 | 0 | **100.0** | 96.6 - 100.0 | 108 | 0 | **100.0** | 96.6 - 100.0 |
| **UCP** | 107 | 2 | **98.1** | 93.4 - 99.8 | 107 | 2 | **98.1** | 93.4 - 99.8 | 107 | 2 | **98.1** | 93.4 - 99.8 |
| **VivaChek** | 99 | 5 | **94.9** | 88.6 - 98.3 | 99 | 4 | **96.0** | 90.0 - 98.9 | 99 | 5 | **94.9** | 88.6 - 98.3 |
| **WondFo** |  |  |  |  |  |  |  |  | 106 | 1 | **99.1** | 94.9 - 100.0 |
| **ELISAs** |  |  |  |  |  |  |  |  |  |  |  |  |
| **Epitope** | 108 | 3 | **97.2** | 92.1 - 99.4 | 108 | 10 | **90.7** | 83.6 - 95.5 | 108 | 11 | **89.8** | 82.5 - 94.8 |
| **In-House** |  |  |  |  |  |  |  |  | 108 | 1 | **99.1** | 94.9 - 100.0 |
| **Percentage of positive specimens from individuals who were positive for non-SARS-CoV-2 viral infections and/or tested negative for SARS-CoV-2 by RT-PCR** |  |  |  |  |  |  |  |  |  |  |  |  |
| **Assay** | **IgM** |  |  |  | **IgG** |  |  |  | **IgM or IgG** |  |  |  |
|  | **Total N** | **positive** | **%** | **95% CI** | **Total N** | **positive** | **%** | **95% CI** | **Total N** | **positive** | **%** | **95% CI** |
| **Immunochromatographic Lateral Flow Assays** |  |  |  |  |  |  |  |  |  |  |  |  |
| **Biomedomics** | 52 | 8 | **15.4** | 6.9 - 28.1 | 52 | 4 | **7.7** | 2.1 - 18.5 | 52 | 11 | **21.2** | 11.1 - 34.7 |
| **Bioperfectus** | 45 | 5 | **11.1** | 3.7 - 24.1 | 45 | 6 | **13.3** | 5.1 - 26.8 | 45 | 8 | **17.8** | 8.0 - 32.1 |
| **DecomBio** | 52 | 5 | **9.6** | 3.2 - 21.0 | 52 | 2 | **3.8** | 0.5 - 13.2 | 52 | 6 | **11.5** | 4.4 - 23.4 |
| **DeepBlue** | 52 | 14 | **26.9** | 15.6 - 41.0 | 52 | 7 | **13.5** | 5.6 - 25.8 | 52 | 14 | **26.9** | 15.6 - 41.0 |
| **Innovita** | 28 | 2 | **7.1** | 0.9 - 23.5 | 28 | 2 | **7.1** | 0.9 - 23.5 | 28 | 3 | **10.7** | 2.3 - 28.2 |
| **Premier** | 52 | 1 | **1.9** | 0.0 - 10.3 | 52 | 1 | **1.9** | 0.0 - 10.3 | 52 | 2 | **3.8** | 0.5 - 13.2 |
| **Sure** | 52 | 0 | **0.0** | 0.0 - 6.8 | 52 | 0 | **0.0** | 0.0 - 6.8 | 52 | 0 | **0.0** | 0.0 - 6.8 |
| **UCP** | 52 | 3 | **5.8** | 1.2 - 15.9 | 52 | 2 | **3.8** | 0.5 - 13.2 | 52 | 3 | **5.8** | 1.2 - 15.9 |
| **VivaChek** | 49 | 4 | **8.2** | 2.3 - 19.6 | 49 | 1 | **2.0** | 0.1 - 10.9 | 49 | 4 | **8.2** | 2.3 - 19.6 |
| **WondFo** |  |  |  |  |  |  |  |  | 41 | 0 | **0.0** | 0.0 - 8.6 |
| **ELISAs** |  |  |  |  |  |  |  |  |  |  |  |  |
| **Epitope** | 52 | 2 | **3.8** | 0.5 - 13.2 | 52 | 8 | **15.4** | 6.9 - 28.1 | 52 | 9 | **17.3** | 8.2 - 30.3 |
| **In-House** |  |  |  |  |  |  |  |  | 52 | 7 | **13.5** | 5.6 - 25.8 |

**Figure 1:**
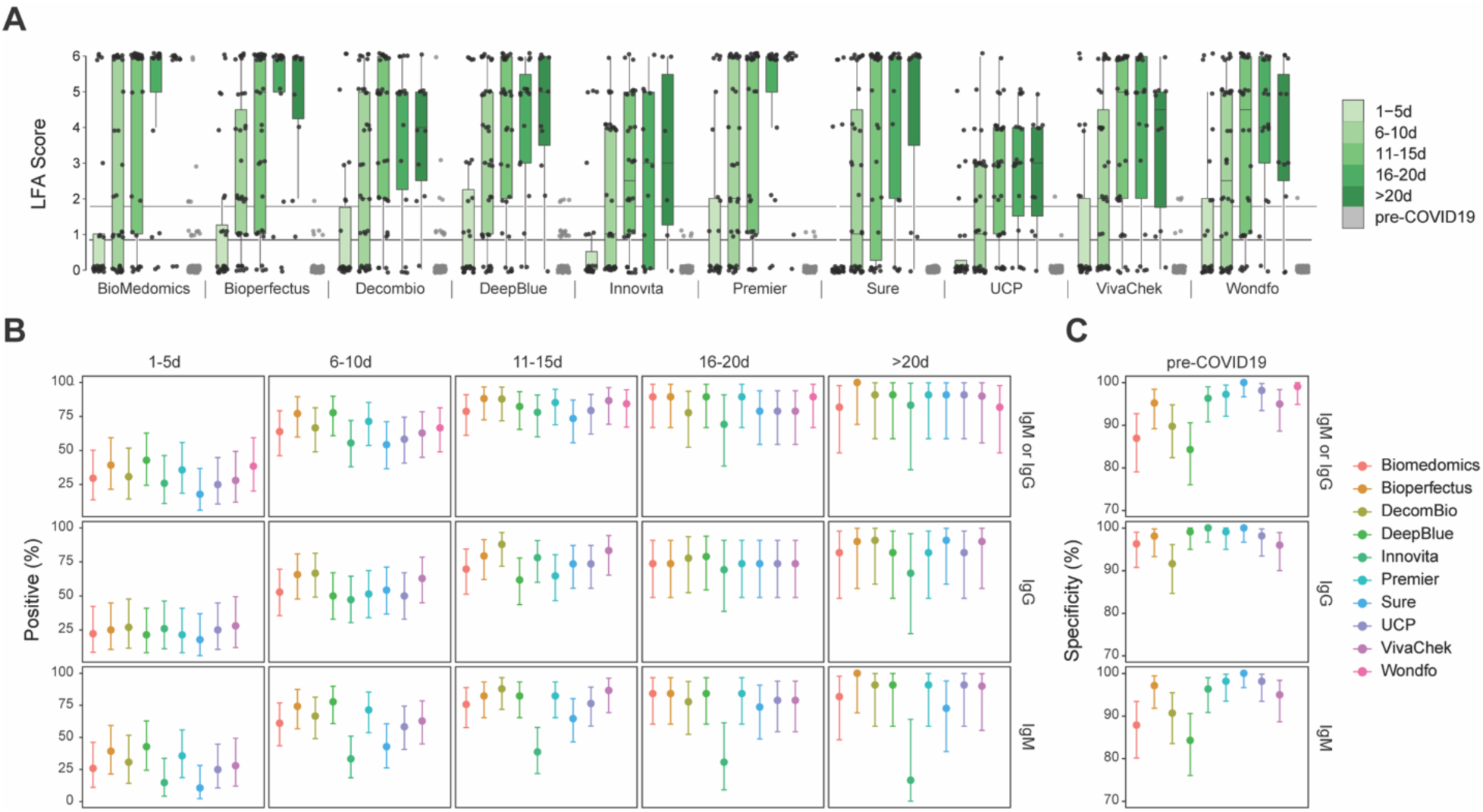
Performance data for immunochromatographic lateral flow assays (LFAs). **A**. The second reader’s score (0-6 based on band intensity) is reported for each assay, binned by time after patient-reported symptom onset. For tests with separate IgG and IgM bands, the higher score is reported. Joint IgM/IgG signal is represented by a single band in Wondfo. The lower, dark grey line refers to the positivity threshold (Score greater than or equal to 1) used in this study. The upper, light grey line refers to an alternative positivity threshold (Score greater than or equal to 2) discussed in the text and eFigure 4. **B**. Percent of SARS-CoV-2 RT-PCR-positive samples testing positive by each LFA are plotted relative to time after patient-reported symptom onset. The “IgM or IgG” category refers to positivity of either isotype. **C**. Specificity is plotted for each test using pre-COVID-19 negative control samples. All error bars signify 95% confidence intervals.

**Figure 2:**
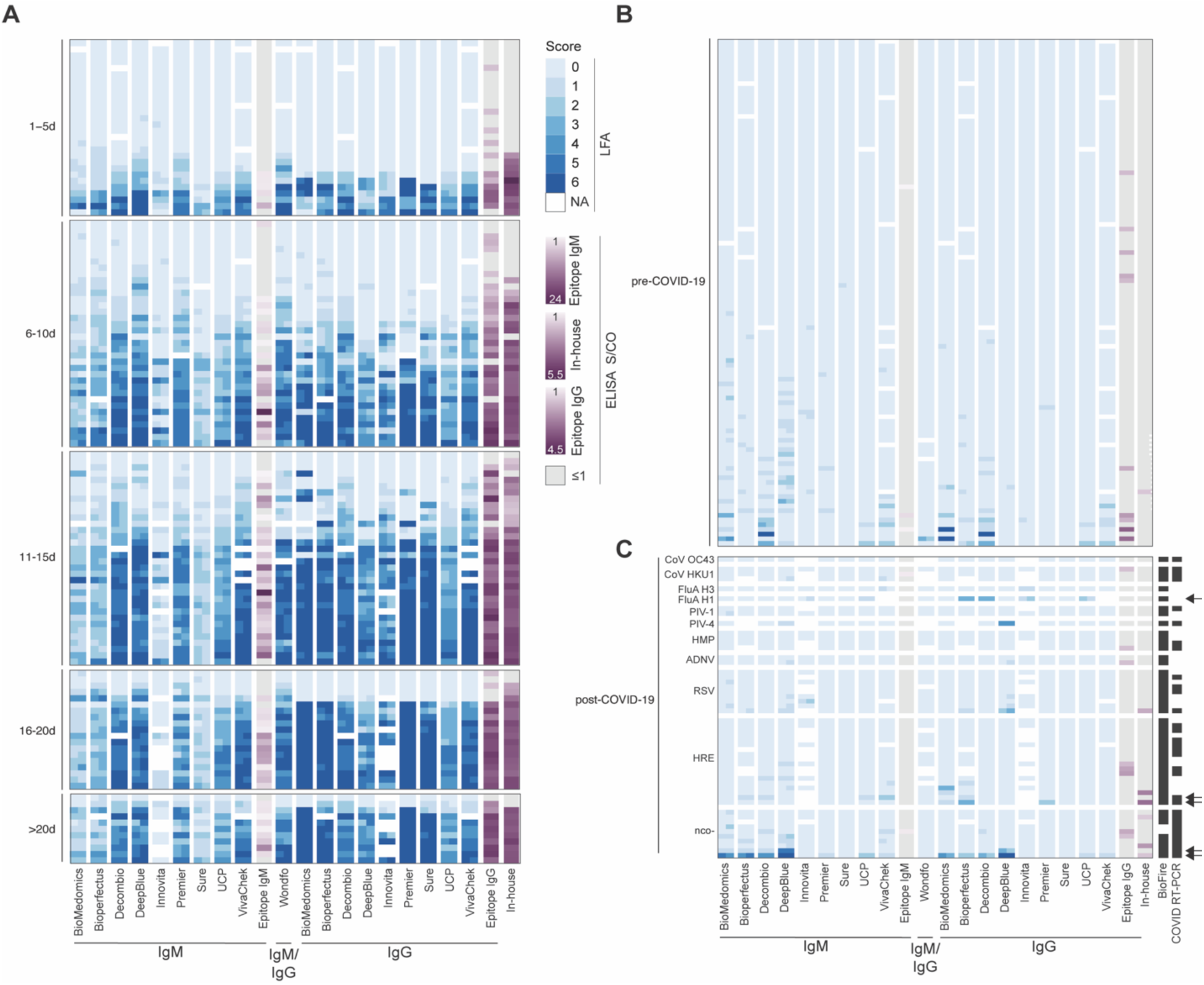
LFA and ELISA values by serological assay. **A**. LFA scores for each of two readers (blue) and mean ELISA Signal/Cutoff Ratio (S/CO, purple) for each specimen are grouped by binned time after patient-reported symptom onset and plotted by assay. White cells indicate samples not run with the corresponding assay. For ELISAs, grey indicates S/CO less than or equal to 1. The same legend applies to Panels B and C. The F(ab’)2 specific secondary antibody used in our in-house ELISA preferentially binds the IgG light chain but has some reactivity for other isotypes (IgM, IgA). **B**. LFA score and ELISA S/CO values are plotted for pre-COVID-19 historical control serum samples to determine assay specificity. **C**. LFA score and ELISA S/CO values are plotted for serum samples obtained from 52 individuals after the emergence of COVID-19 (post-COVID-19), some of which received Biofire FilmArray (BioFire Diagnostics, Salt Lake City, UT) and/or SARS-CoV-2 RT-PCR testing (all negative) as indicated (black cells) in the appropriate columns. Arrows highlight specimens from five individuals with moderate to strong band intensity further discussed in the text. Specimens are grouped by positive testing for Coronavirus HKU1 (CoV HKU1), Coronavirus OC43 (CoV OC43), Influenza A Virus A/H3 (FluA H3), Influenza A Virus A/H1 2009 (FluA H1), Parainfluenza Type 1 Virus (PIV-1), Parainfluenza Type 4 Virus (PIV-4), Human Metapneumovirus (HMP), Adenovirus (ADNV), Respiratory Syncytial Virus (RSV), Human Rhinovirus/Enterovirus (HRE), or negative testing for SARS-CoV-2 and other viruses (nco-).

We observed a trend towards higher percent positivity by LFA for patients admitted to ICU compared to those with milder disease, but the specimen numbers per time interval were low, limiting statistical power (eFigure 3).

Test specificity in pre-COVID-19 samples ranged from 84.3%-100.0%, with 39 samples demonstrating false positive results by at least one LFA (Table 2 and Figure 2B). Of the false positive results, 61.5% (24/39) had a weak intensity score (1). Intensity scores of 2-3 were seen in 30.8% (12/39) and scores of 4-6 were seen in 7.7% (3/39).

We evaluated the tradeoff between percent positivity and specificity as a function of LFA reader score. Changing the positive LFA threshold from 1 to 2 decreased the mean overall percent positivity across tests from 67.2% (range: 57.9%-75.4%) to 57.8% (range: 44.7%-65.6%) and increased the average specificity from 94.2% (range: 84.3%-100.0%) to 98.1% (range: 94.4%-100.0%) (eFigure 4). An independent study at MGH compared three LFAs, of which BioMedomics was also assessed in the current study (eTable 3). Overall, both studies showed a trend for increased detection of SARS-CoV-2 specific antibodies with increased time from symptom onset. However, the MGH study displayed increased specificity with lower percent positivity at early timepoints after symptom onset. MGH positivity thresholds were set higher to prioritize test specificity (eFigure 4B-C).

A set of specimens obtained during the COVID-19 outbreak that had negative SARS-CoV-2 RT-PCR testing and/or alternative respiratory pathogen diagnoses demonstrated higher numbers of positive results compared to the pre-COVID-19 sample set (Figure 2C). Five specimens had positives results by >3 tests, all with respiratory symptoms and concurrent negative SARS-CoV-2 RT-PCR testing (Figure 2C, arrows). One patient was positive on 8 different tests including the in-house ELISA. In this limited panel, no consistent pattern of cross-reactivity was identified in samples from individuals with non-SARS-CoV-2 respiratory viruses, including 2 strains of seasonal coronavirus (1 coronavirus OC43, 3 coronavirus HKU1).

Agreement between results of LFAs with those of IgG and IgM Epitope ELISAs ranged from 75.7%-85.6%, while agreement with the in-house ELISA ranged from 83.5%-94.8% (Figure 3A). LFA band intensity scores showed a direct correlation with ELISA S/CO values (Figure 3B).

**Figure 3:**
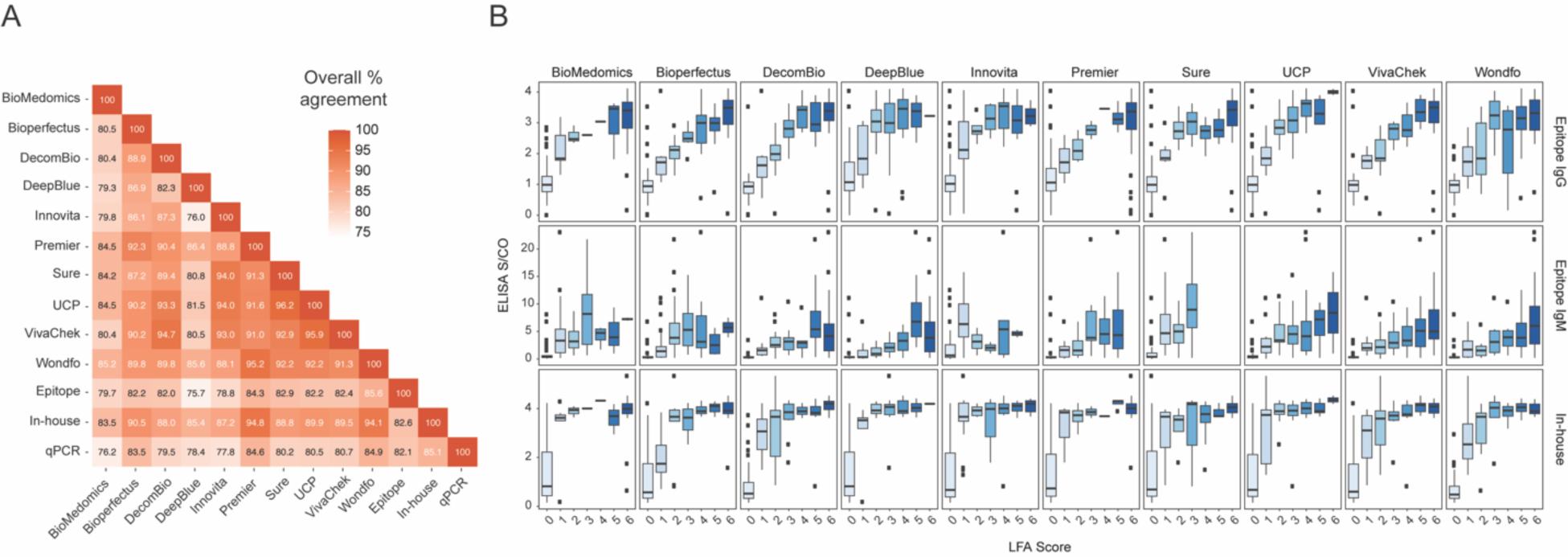
Agreement of serological assays for SARS-CoV-2. **A**. Percent agreement is plotted across all assay combinations, and values signify the binomial regression of the two assays across all tests. Samples were labeled “positive” if any one isotype was detected (LFA score ≥ 1, S/CO > 1) for each assay. **B**. IgM or IgG LFA scores for each assay are compared to Signal/Cutoff Ratios (S/CO) from three different ELISAs for all SARS-CoV-2 RT-PCR-positive samples. Joint IgM/IgG signal is represented by a single band in Wondfo, so data were plotted as IgM or IgG depending on ELISA comparison. The F(ab’)2 specific secondary antibody used in our in-house ELISA preferentially binds the IgG light chain but contains some reactivity for other isotypes (IgM, IgA). Error bars signify 95% confidence intervals.

## DISCUSSION

This study describes test performance for 12 COVID-19 serology assays on a panel of 128 samples from 79 individuals with PCR-confirmed SARS-CoV-2 infection and 108 pre-COVID-19 specimens. For each test, we quantified detection of IgM and/or IgG antibodies by time period from onset of symptoms and assessed specificity and cross-reactivity. We hope these data will inform the medical community, public health efforts, and governmental institutions considering SARS-CoV-2 serological testing. This study also seeks to provide feedback to manufacturers about areas of success and necessary improvement. There is no “gold standard” to identify true seropositive blood samples. The extent and time-course of antibody development are not fully understood as yet, and may vary between different populations, even among RT-PCR-confirmed cases.

We focused on comparisons of percent positivity by time interval, rather than reporting the “sensitivity” of each assay. As expected, percent positivity rose with time after symptom onset.^5,6,21-24^ High rates of positive results were not reached until at least 2 weeks into clinical illness; diagnosis at time of symptom onset thus remains dependent on viral detection methods. The assays showed a trend to higher positive rates within time intervals for more severe disease, but this finding should be interpreted with caution, due to the limited data from ambulatory cases. The majority of samples >20 days post-symptom onset had detectable anti-SARS-CoV-2 antibodies, suggesting good to excellent sensitivity for all evaluated tests in hospitalized patients three or more weeks into their disease course. However, well-powered studies testing ambulatory or asymptomatic individuals, including performance with capillary blood, will be essential to guide appropriate use of serology.

Our data demonstrate specificity greater than 95% for the majority of tests evaluated and >99% for 2 LFAs (Wondfo, Sure Biotech) and the in-house ELISA (adapted from Amanat et al, 18 2020). We observed moderate-to-strong positive bands in several pre-COVID-19 blood donor specimens, some of them positive by multiple assays, suggesting the possibility of non-specific binding of plasma proteins, non-specific antibodies, or cross-reactivity with other viruses. Three of the pre-COVID-19 specimens (2.8%) were scored positive by more than three assays. Intriguingly, the fraction of positive tests was higher in a set of recent specimens obtained during the COVID-19 outbreak from individuals undergoing respiratory viral workup, many with negative SARS-CoV-2 RT-PCR. Five of these (9.6%) had positive results by more than three assays, without relation to a specific viral pathogen, suggesting non-specific reactivity and/or missed COVID-19 diagnoses. One specimen was positive by 8 of 12 assays, including the in-house ELISA. The patient was >90 years old and presented with altered mental status, fever, and ground glass opacities on chest radiological imaging. SARS-CoV-2 RT-PCR was negative and ancillary laboratory testing suggested a urinary tract infection. This case could represent COVID-19 not detected by RT-PCR, reinforcing the importance of caution in interpreting negative molecular results as ruling out the infection. Appropriate algorithms for serology testing, including confirmatory or reflexive testing, have yet to be determined. These algorithms will be affected by test performance characteristics and prevalence of disease, as well pretest probability of infection.

Importantly, we still do not know the extent to which positive results by serology reflect a protective immune response.^25^ Future functional studies are critical to determine whether specific antibody responses predict virus neutralization and protection against re-infection. Until this is established, conventional antibody assays should not be used as predictors of future infection risk.

High specificity testing is crucial in low-prevalence settings. One approach to increase specificity would employ confirmatory testing with an independent assay (perhaps recognizing a distinct epitope or antigen). Our comparison of UCSF and MGH data suggests that reclassifying faint bands as negative or inconclusive changes test performance characteristics by increasing specificity, albeit at the expense of sensitivity. However, the subjectivity of calling faint bands by individual readers may be difficult to standardize without specific control materials, operator training, and/or objective methods of analyzing LFAs. In the clinical setting, these parameters and protocols should be independently assessed and validated by clinical laboratories for operation under the Clinical Laboratory Improvement Amendments (CLIA).^26^

Our study also reinforces the need for assay validation using standardized sample sets with: 1) known positives from individuals with a range of clinical presentations at multiple time points after onset of symptoms, 2) pre-COVID-19 outbreak samples for specificity, and 3) samples from individuals with other viral and inflammatory illnesses as cross-reactivity controls. Coordinated efforts to validate and ensure widespread availability of such standardized sample sets would facilitate effective utilization. Serology test performance data will be available on a dedicated website (https://covidtestingproject.org). Current and future studies by our group and others will provide an essential evidence base to guide serological testing during the COVID-19 pandemic.

## Data Availability

All data used in the study are available in an interactive format at https://covidtestingproject.org/x

https://covidtestingproject.org/

## Acknowledgements

We thank all members of the Marson lab and the Hsu lab, Peter Kim, Scott Boyd, Joe DeRisi, Steve Quake, Bryan Greenhouse, Christina Tato, Jennifer Doudna, Fyodor Urnov, David Friedberg, David Neeleman, John Hering, Cindy Cheng, Neal Khosla, Matt Krisiloff, Lachy Groom, Chenling Xu, Dave Fontenot, Jim Karkanias, Gajus Worthington, Bill Burkholder, Charlie Craik, XPrize Pandemic Alliance, Warris Bokhari, Zem Joaquin, Siavash Sarlati, Scott Nesbit, William Poe, Sam Broder, Verily, Charlie Kim, Aleksandra Kijac, Marc Solit and the Coronavirus Standards Working Group, Diane Havlir, Joanne Engel, Peter Farley, Jeff MacGregor, Kimberly Hou, Bob Sanders, Sarah Yang, and Sean Parker. We thank Yagahira Elizabeth Castro-Sesquen for sharing her semi-quantitative LFA scale, which was adapted for use in our current study. The work was supported by gifts from Anthem Blue Cross Blue Shield, the Chan Zuckerberg Biohub, and anonymous philanthropy. We thank the following sources for donation of test kits: the manufacturers of Bioperfectus, Decombio, Sure-Bio, UCP Biosciences; David Friedberg; John Hering; Henry Schein (Melville, NY). The Wilson Lab has received support from the Rachleff Family Foundation. The Hsu lab has received support from S. Altman, V. and N. Khosla, D. and S. Deb, the Curci Foundation, and Emergent Ventures. P.D.H. holds a Deb Faculty Fellowship from the UC Berkeley College of Engineering and is the recipient of the Rainwater Foundation Prize for Innovative Early-Career Scientist. The Marson lab has received gifts from J. Aronov, G. Hoskin, K. Jordan, B. Bakar, the Caufield family and funds from the Innovative Genomics Institute (IGI), the Northern California JDRF Center of Excellence and the Parker Institute for Cancer Immunotherapy (PICI). We thank the National Institutes of Health for their support (J.D.W. R38HL143581; A.E.G. F30AI150061; D.N.N. L40 AI140341; S.P.B. NHLBI R38HL143581; T.A.M. 1F30HD093116; D.W NIH 1F31NS106868-01; J.G.C. R01 AI40098; E.T.R., R.C.C. CDC U01CK000490; MSTP students supported by T32GM007618). R.Y. was supported by an AP Giannini Postdoctoral Fellowship. J.A.S. was supported by the Larry L. Hillblom Foundation (2019-D-006-FEL). A.M. holds a Career Award for Medical Scientists from the Burroughs Wellcome Fund, is an investigator at the Chan-Zuckerberg Biohub and is a recipient of The Cancer Research Institute (CRI) Lloyd J. Old STAR grant.

## Competing Interests

This work was supported by gifts from Anthem Blue Cross Blue Shield, the Chan Zuckerberg Biohub, and anonymous philanthropy. C.Y.C. is the director of the UCSF-Abbott Viral Diagnostics and Discovery Center, receives research support funding from Abbott Laboratories and is on the Scientific Advisory Board of Mammoth Biosciences, Inc. C. J. Y. is cofounder of DropPrint Genomics and serves as an advisor to them. M.S.A. holds stock in Medtronic and Merck. P.D.H. is a cofounder of Spotlight Therapeutics and serves on the board of directors and scientific advisory board, and is an advisor to Serotiny. P.D.H. holds stock in Spotlight Therapeutics and Editas Medicine. A.M. is a cofounder of Spotlight Therapeutics and Arsenal Biosciences and serves on their boards of directors and scientific advisory boards. A.M. has served as an advisor to Juno Therapeutics, was a member of the scientific advisory board at PACT Pharma, and was an advisor to Trizell. A.M. owns stock in Arsenal Biosciences, Spotlight Therapeutics and PACT Pharma. RY owns stock in Abbvie, Bluebird Bio, Bristol Myers Squibb, Cara Therapeutics, Editas Medicine, Esperion, and Gilead Sciences. Unrelated to this current work, the Marson lab has received sponsored research support from Juno Therapeutics, Epinomics and Sanofi, and a gift from Gilead.

